# Retrieval-Augmented Large Language Models for Clinically Aligned Adverse Event Coding in Acute Myeloid Leukemia Clinical Trials

**DOI:** 10.64898/2026.08.17.26360282

**Authors:** Naghme Dashti, Martin M. K. Schneider, Jan-Niklas Eckardt, Frank Fiebig, Dirk Schweigler, Sophia Büttner, Moritz Middeke, Martin Bornhäuser, Christoph Röllig, Jakob Nikolas Kather, Isabella C. Wiest

## Abstract

**Background:** Adverse event (AE) coding is essential for safety monitoring in oncology clinical trials, particularly in acute myeloid leukemia (AML), where intensive therapies are associated with frequent and heterogeneous toxicities requiring standardized MedDRA (Medical Dictionary for Regulatory Activities) coding. However, manual Low-Level Term (LLT) assignment remains labor-intensive, subjective, and difficult to scale. Although large language models (LLMs) have emerged as promising decision-support tools for automated coding, unguided zero-shot generation remains insufficient for reliable fine-grained MedDRA coding.

**Objective:** To develop and evaluate a retrieval-augmented reasoning pipeline for clinically aligned LLT-level MedDRA coding of free-text adverse events from prospective AML clinical trials.

**Methods:** We implemented a retrieval-augmented reasoning pipeline inspired by the retrieval-augmented generation (RAG) paradigm using LLaMA-3.3-70B-Instruct as the primary backbone and benchmarked the framework across multiple open instruction-tuned LLMs. Dense semantic retrieval first generated a constrained top-100 LLT candidate set for each AE, followed by structured LLM reasoning to select a single best-matching LLT and deterministic mapping to Preferred Term (PT) and System Organ Class (SOC) levels. The pipeline was evaluated retrospectively on AE datasets from three prospective AML clinical trials (MOSAIC, DELTA, and DaunoDouble) with automated LLT/PT/SOC metrics and expert-assessed Clinical Correctness Rate (CCR).

**Results:** Clinical expert review showed high clinical acceptability of the RAG pipeline across datasets (91-97%). Under automated evaluation, the pipeline achieved LLT exact accuracy of 50-58%, PT accuracy of 78-85%, and SOC accuracy of 90-93%. Zero-shot generation and random candidate selection performed substantially worse. Semantic retrieval more often included the coder-assigned LLT among the candidate terms available to the model than retrieval based on lexical similarity. Multi-model benchmarking showed that backbone choice mainly affected LLT exact agreement, whereas PT and SOC performance remained comparatively stable.

**Conclusions:** Retrieval-augmented reasoning supports clinically aligned MedDRA coding of free-text adverse events under realistic candidate constraints in AML clinical trials. Evaluation across three AML clinical trials showed that strict LLT-level string agreement underestimated clinical appropriateness, highlighting the importance of combining hierarchical evaluation metrics with clinical expert validation for AI-assisted MedDRA coding in hematology trials.

## INTRODUCTION

Adverse event (AE) documentation is a core element of Good Clinical Practice and a key regulatory requirement for clinical trials and pharmacovigilance. In clinical research, AEs are defined as any unfavorable and unintended sign, symptom, or disease temporally associated with the use of a medicinal product, irrespective of causality (1). Although AEs do not necessarily indicate treatment-related harm, their consistent documentation is essential for evaluating drug safety profiles, informing regulatory decisions, and supporting ongoing patient protection efforts (1,2). Despite this importance, AE descriptions are frequently heterogeneous, incomplete, or inconsistently reported across clinical trials, hospital registries, and real-world data sources, limiting the reliability of large-scale safety surveillance (3,4).

Accurate and standardized AE classification is enabled globally through the Medical Dictionary for Regulatory Activities (MedDRA), an internationally hierarchical terminology containing more than 80,000 Low Level Terms (LLTs) mapped to clinically meaningful Preferred Terms (PTs) and System Organ Classes (SOCs) (5). MedDRA coding is required for clinical trial submissions, pharmacovigilance case processing, safety signal detection, and post-marketing surveillance (1,5,6). Free-text AE-to-MedDRA mapping is still performed manually; clinical coders and safety specialists must interpret free-text AE narratives, often ambiguous, variably phrased, or institution-specific, and identify the best-matching LLT or PT (3,4). This process is labor-intensive, cognitively demanding, and prone to inter-rater variability, particularly in large clinical trials, such as oncology studies, where thousands of AEs may be recorded (3). To address these challenges, AI-assisted coding using large language models (LLMs) has emerged as a promising decision-support approach to support clinical coders, reduce manual burden, and facilitate scalable clinical coding workflows (7,8).

Acute myeloid leukemia (AML) clinical trials represent a particularly relevant setting for adverse event documentation and standardized MedDRA coding. As therapeutic strategies for hematologic malignancies become increasingly complex due to the advent of targeted agents, comprehensive and standardized adverse event assessment has become increasingly important for reliable safety evaluation, regulatory reporting, and comparison across clinical studies (9). These evolving requirements underscore the importance of robust and reproducible approaches for MedDRA-based adverse event coding in hematology clinical trials.

Early research into automated AE coding employed rule-based natural language processing (NLP), dictionary lookup systems, and traditional machine-learning models (4,10). While these approaches provided partial automation, they struggled with synonymy, linguistic variability, and context-dependent interpretation inherent in clinical narratives (4,7,10). Recent advances in LLMs, such as GPT-4 and domain-adapted clinical transformers, have substantially improved the ability to interpret unstructured medical text and perform coding-like tasks such as symptom normalization, clinical concept extraction, and classification (11–13). However, most prior studies have evaluated coding only at higher MedDRA levels (e.g., PT or HLT-level) rather than at the LLT level, where the most fine-grained clinical information is represented; while PT-level coding may be sufficient for aggregate analyses, it can obscure clinically relevant detail present in free-text AE descriptions. Moreover, few studies have systematically assessed LLM performance under clinical review (8,14), a critical requirement for safety-relevant applications. Relatedly, prior work has shown that zero-shot LLMs alone are insufficient for reliable, fine-grained medical coding, particularly in highly granular terminologies such as MedDRA, where multiple LLTs may represent closely related but clinically distinct concepts (14,15). Accordingly, zero-shot LLM performance primarily serves as a lower-bound reference rather than a practical coding solution.

Beyond improving coding accuracy, there remains a need for evaluation frameworks that assess whether LLM-assisted coding is clinically appropriate for safety-critical applications. To address these gaps, we present a retrieval-augmented LLM pipeline for automated MedDRA coding of free-text AEs. Our approach combines dense semantic retrieval with explicit structured reasoning using a state-of-the-art LLM to generate a single interpretable LLT prediction for each AE.

Using three independent AML prospective clinical trial datasets, we evaluate the ability of this end-to-end pipeline to generate clinically appropriate LLT predictions from free-text AE descriptions under realistic MedDRA coding conditions. For interpretive context, performance is benchmarked across MedDRA’s hierarchical levels (LLT, PT, SOC) and is compared against zero-shot generation as well as random and lexical candidate construction strategies. In addition, clinical correctness is assessed through expert review, an evaluation dimension rarely examined in prior work. Together, this work provides an interpretable and clinically grounded framework for evaluating AI-assisted MedDRA coding in AML clinical trials and supports future integration of LLM-based decision-support systems into hematology research and pharmacovigilance workflows.

## METHODS

### Study Design and Data Sources

We conducted a retrospective evaluation of LLM-based MedDRA coding using free-text AE documentation extracted from three prospective controlled clinical trials in patients with acute myeloid leukemia (AML), under conditions reflecting routine hematology clinical trial coding and pharmacovigilance workflows. The primary objective was to assess whether a retrieval-augmented reasoning pipeline inspired by the retrieval-augmented generation (RAG) paradigm could generate clinically appropriate MedDRA codes from free-text adverse event descriptions collected in prospective AML clinical trials, despite their heterogeneous phrasing and lexical ambiguity. The evaluation used AE datasets from MOSAIC (NCT04385290), DELTA (NCT02446145) (16), and DaunoDouble (NCT02140242) (17), which were collected until August 2024 (See Table 1). After harmonization and removal of duplicate AE text entries, the final evaluation datasets contained 179 unique AE descriptions from MOSAIC, 226 from DELTA, and 443 from DaunoDouble. These counts refer to the deduplicated AE descriptions used for this methodological evaluation and should not be interpreted as the total number of adverse events reported in the original clinical trials. Only anonymized retrospective data were analyzed. All studies were conducted in accordance with the revised Declaration of Helsinki (18) and had prior institutional review board approval. Each dataset contained verbatim AE descriptions paired with coder-assigned MedDRA LLTs and PTs (Coded-LLT, Coded-SOC) from MedDRA v25.0, which served as the human reference for automated evaluation. LLTs were deterministically linked to their corresponding PT and System Organ Class (SOC) terms. Figure 1 illustrates the mapping from free-text AE descriptions to LLT, PT, and SOC categories. This structure permitted analysis of both lexical alignment at the LLT level and broader clinical categorization.

**Table 1.** Dataset characteristics, including AE counts, unique LLTs, PTs, SOCs, and the five most frequent SOC categories per dataset. Abbreviations: AE = Adverse Event; LLT = Low Level Term; PT = Preferred Term; SOC = System Organ Class.

| Dataset | #AEs | Unique LLTs | Unique PTs | Unique SOCs | Top SOCs (count) |
| --- | --- | --- | --- | --- | --- |
| DaunoDouble | 443 | 199 | 159 | 21 | Infections and infestations (154); Blood and lymphatic system disorders (66); General disorders and administration site conditions (62); Gastrointestinal disorders (47); Nervous system disorders (42) |
| DELTA | 226 | 115 | 100 | 19 | Infections and infestations (64); Vascular disorders (26); Gastrointestinal disorders (23); Nervous system disorders (22); Skin and subcutaneous tissue disorders (21) |
| MOSAIC | 179 | 82 | 75 | 20 | Vascular disorders (23); Blood and lymphatic system disorders (19); Infections and infestations (17); Gastrointestinal disorders (15); General disorders and administration site conditions (14) |

**Figure 1.**
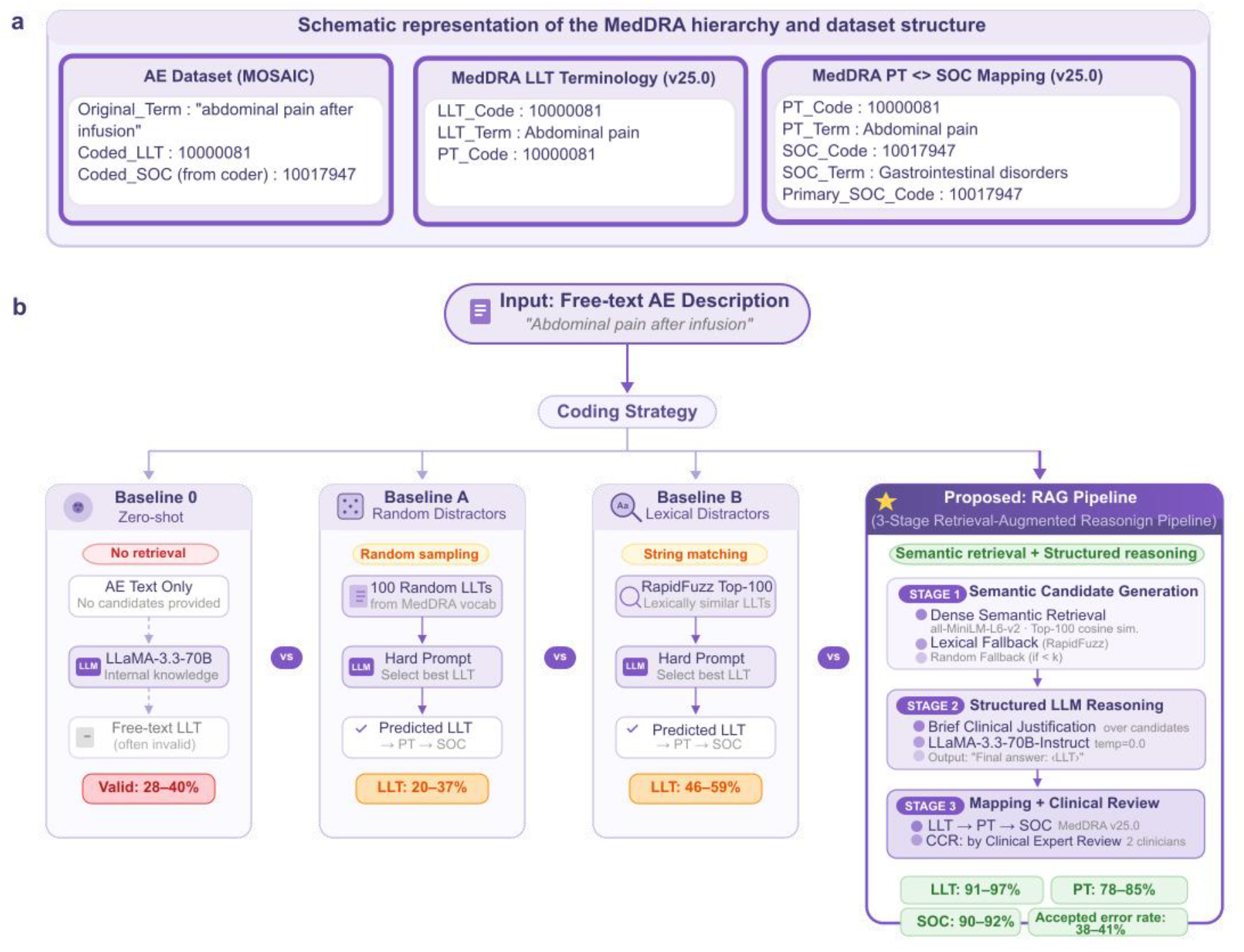
Overview of the MedDRA hierarchy, dataset structure, and evaluation framework for MedDRA-LLM. a) Schematic representation of the MedDRA hierarchy and dataset structure. Free-text adverse event (AE) descriptions from three prospective AML clinical trial datasets are manually coded to MedDRA Low Level Terms (LLTs), which are deterministically linked to Preferred Terms (PTs) and System Organ Classes (SOCs) according to MedDRA v25.0. This hierarchical structure enables evaluation at multiple abstraction levels (LLT, PT, SOC) b) Comparison of MedDRA coding strategies. Three prompt-based baselines are evaluated against the proposed retrieval-augmented reasoning pipeline. Baseline 0 performs zero-shot LLT generation without candidate retrieval. Baseline A uses randomly sampled LLT candidates, while Baseline B uses lexically similar candidates obtained with RapidFuzz string matching. The proposed MedDRA-LLM pipeline consists of three stages: (1) semantic candidate generation, where dense semantic retrieval (MiniLM embeddings) serves as the primary retrieval mechanism for top-k LLT exposure, with lexical fallback and random completion used only when dense retrieval is unavailable or does not meet predefined retrieval-confidence criteria; (2) structured LLM reasoning, where an instruction-tuned LLaMA-3.3-70B model first generates a brief structured justification over the retrieved LLT candidates before producing a constrained final LLT prediction; and (3) hierarchical mapping and clinical validation, where predicted LLTs are deterministically mapped to PT and SOC levels and a subset of predictions undergo clinical expert review to assess the clinical validity of model predictions (Clinical Correctness Rate, CCR). Performance is reported using LLT-exact accuracy, PT/SOC agreement, and clinical validation metrics.

### Dataset Construction and Preprocessing

Adverse event descriptions and their corresponding MedDRA labels were extracted from structured trial files and harmonized across datasets. All LLT and PT terms from MedDRA v25.0 were compiled into structured CSV and JSON resources containing codes and hierarchical mappings. To support semantic retrieval, dense vector representations of all LLTs were generated using the all-MiniLM-L6-v2 sentence transformer (19), which produces 384-dimensional embeddings optimized for semantic similarity. These embeddings were stored in a persistent JSON knowledge base and indexed for candidate generation. Details of retrieval and fallback logic are described in the System Architecture section below.

### System Architecture: MedDRA-LLM Pipeline

The MedDRA-LLM system was implemented as a retrieval-augmented reasoning pipeline consisting of three sequential stages.

#### Stage 1

Semantic Candidate Generation. A semantic-first retrieval mechanism was used to construct a concise candidate set for each adverse event description. First, dense semantic retrieval identified the top-k (k=100) most similar LLTs based on cosine similarity between AE and LLT embeddings. Dense retrieval served as the primary candidate generation strategy. In rare cases where AE embeddings were unavailable or dense retrieval did not meet predefined retrieval-confidence criteria, a RapidFuzz token-set similarity search was used as lexical fallback to supplement the candidate list (20). If fewer than k candidates were available after lexical fallback, a small random subset of LLTs was appended to preserve a fixed candidate list size. This process constrained the search space for downstream reasoning but did not determine the final prediction.

#### Stage 2

Structured LLM Reasoning. Candidate lists were converted into structured prompts and evaluated using “LLaMA-3.3-70B-Instruct”, an instruction-tuned large language model deployed locally (21). Each prompt required the model to first generate a brief structured clinical justification before producing a single machine-parsable output in the form “Final answer: <LLT_TERM>“. Only this final line was extracted as the predicted LLT. Unlike the prompt-only baselines, which directly select an LLT from the provided candidate list, this stage combines structured LLM reasoning with semantically retrieved LLT candidates before producing the final LLT prediction. This design ensured reproducibility, prevented free-text drift, enabled automated hierarchical mapping, and was used strictly in inference mode without fine-tuning. Although the primary experiments used LLaMA-3.3-70B-Instruct, additional benchmarking was performed with several alternative instruction-tuned open-source LLMs to assess whether performance was sensitive to the choice of model under the same retrieval, prompting, and evaluation setup.

#### Stage 3

Deterministic Mapping. Predicted LLTs were mapped deterministically to their associated Preferred Term (PT) and System Organ Class (SOC) using MedDRA v25.0 hierarchical relationships (LLT→PT→SOC). Primary SOC rules (based on Primary_SOC_Code when applicable) were applied to provide consistent SOC-level evaluation, enabling assessment across multiple clinical abstraction levels (LLT, PT, SOC). Therefore, differences between the RAG pipeline and the baselines reflect both improved candidate exposure through semantic retrieval and the use of structured reasoning for final selection.

To support interactive evaluation and simulate real-world usage, we implemented a lightweight user interface that allows batch upload of AE descriptions (e.g., via spreadsheet files), automated MedDRA coding using the proposed pipeline, and optional user-driven clinical validation of predicted LLTs.

### Baseline Methods for Comparison

To contextualize the contribution of the RAG-based reasoning pipeline, we implemented three prompt-only baselines that represent increasingly informed approaches to MedDRA LLT selection, ranging from no grounding at all (zero-shot) to random and lexical candidate construction. These baselines define a prompt-only performance envelope for automated AE-to-LLT mapping. Unlike the RAG pipeline, which incorporates structured reasoning, both Baseline A and Baseline B operate as prompt-only selection mechanisms. In these baselines, the model is instructed to directly select the most appropriate LLT term from the provided candidate list without performing explicit reasoning.

*Baseline 0)* Zero-shot LLT Generation. In this lower-bound condition, the LLM received only the free-text AE description and was asked to output a single MedDRA LLT term, without access to any candidate list or dictionary entries. Model outputs were then matched against the MedDRA LLT dictionary to determine (i) whether the prediction corresponded to a valid LLT, and measured (ii) LLT, PT, and SOC-level accuracy relative to the human baseline. This baseline addresses whether a general LLM, without any code grounding, can perform MedDRA coding reliably (expected to be poor).

*Baseline A)* Hard Prompt with Randomly Sampled LLT Candidates. The candidate set consisted of 100 randomly sampled LLTs (k = 100) from the MedDRA vocabulary. The coder-assigned LLT was neither manually inserted nor explicitly excluded; thus, this baseline represents an uninformed random candidate list drawn from the terminology. This condition captures the inherent difficulty of selecting the correct term in the absence of any pre-filtering, effectively simulating an uninformed search across a large and heterogeneous candidate space.

*Baseline B*) Hard Prompt with Lexically Retrieved Candidates. For this baseline, the candidate set was constructed by selecting the top-k (k=100) LLTs with the highest string-level similarity to the AE description using RapidFuzz. The coder-assigned LLT was not manually inserted or excluded; if it appeared in the candidate list, it did so naturally through lexical retrieval. This approach reflects heuristic workflows where lexical similarity is used as a pre-filter for candidate exposure.

The RAG pipeline replaces uninformed random candidate construction with dense semantic retrieval (all-MiniLM-L6-v2) as the primary candidate generation strategy and integrates structured reasoning. In rare low-confidence or embedding-unavailable cases, lexical fallback retrieval is used to preserve robust candidate exposure. Unlike the baselines, the RAG condition combines constrained candidate exposure with explicit structured reasoning for final selection.

All baseline methods and the RAG pipeline were executed independently three times per dataset using different random seeds. For all LLT, PT, and SOC-level evaluations, we report mean and standard deviation across seeds to ensure robustness, reproducibility, and fair comparison between methods (Figure 2 and Figure 4). The full prompt templates used for all baselines and the RAG pipeline are provided in the Supplementary Materials.

**Figure 2.**
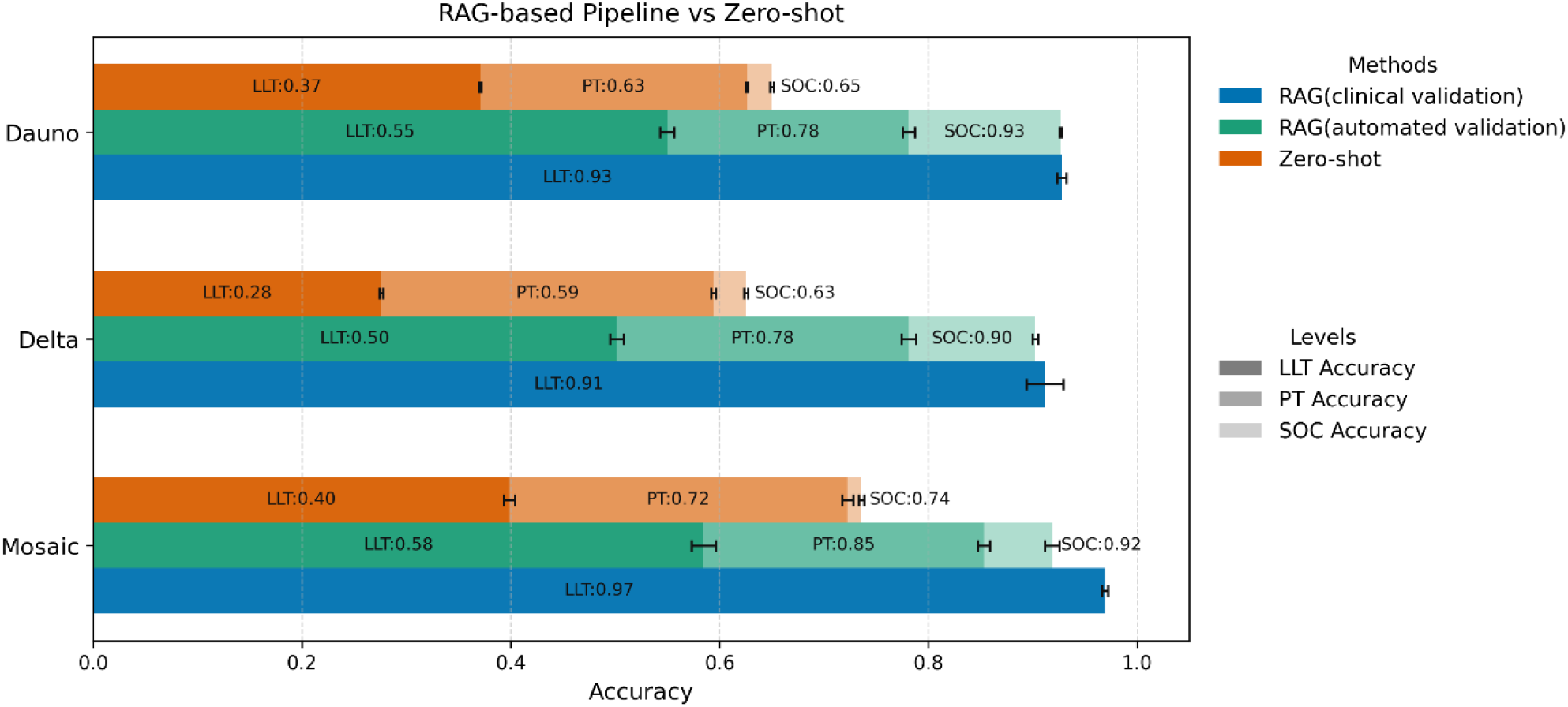
Performance comparison of the RAG-based pipeline and a zero-shot baseline across MedDRA hierarchy levels. Low Level Term (LLT), Preferred Term (PT), and System Organ Class (SOC) accuracies are shown for the three AML clinical trial datasets: MOSAIC, DELTA, and DaunoDouble datasets. Bars represent mean accuracies aggregated across three random seeds for the RAG-based pipeline (automated validation) and the zero-shot baseline; error bars indicate standard deviation. For the RAG-based pipeline, clinical validation was assessed separately at the LLT level through clinical expert review and is shown as the clinically validated RAG pipeline. PT and SOC accuracies are computed via hierarchical mapping from the predicted LLT, with direct comparison with coder-provided PT and SOC reference labels. **Abbreviations:** LLT = Low Level Term; PT = Preferred Term; SOC = System Organ Class; RAG = Retrieval-Augmented Generation; here used to refer to the RAG-inspired retrieval-augmented reasoning pipeline.

### Evaluation Metrics

Model performance was evaluated using automated lexical and hierarchical metrics together with clinical validation. Automated evaluation included LLT exact accuracy, fuzzy LLT accuracy, PT and SOC agreement, and macro-averaged precision, recall, and F1. To assess the clinical validity of model predictions beyond automated performance metrics, a subset of predictions underwent clinical expert review, summarized as the Clinical Correctness Rate (CCR). Two independent clinicians reviewed model-predicted LLTs for regulatory acceptability, and disagreements were resolved by a third clinician. Formal metric definitions and detailed evaluation procedures are provided in the Supplementary Materials.

## RESULTS

We evaluated MedDRA LLT prediction using three prompt-only baselines (Baseline 0: zero-shot; Baseline A: randomly sampled LLT candidates; Baseline B: lexically retrieved candidates) and the RAG pipeline. Performance was assessed through automated comparison with coder-provided reference labels and clinical expert validation. Unless otherwise stated, results are reported as mean ± standard deviation across three random seeds.

### Clinical expert validation reveals higher clinical utility than automated LLT agreement

The RAG-based pipeline supported clinically aligned AE coding across all three AML clinical trial datasets. Clinical expert review showed high clinical acceptability (91-97%), whereas automated LLT exact agreement with coder-provided labels was lower (50-58%). Despite the lower LLT agreement, PT and SOC agreement was retained (78-85% and 90-93%, respectively). In the zero-shot setting (Baseline 0), the LLM frequently produced outputs which did not match the human coder-provided MedDRA LLT labels. Specifically, zero-shot LLT exact accuracy was 40% in MOSAIC, 28% in DELTA, and 37% in DaunoDouble, indicating that most zero-shot outputs did not exactly match the coder-provided reference LLT.

Clinical validation shows that automated comparison with the coder-provided reference labels underestimates expert-assessed clinical appropriateness. Automated LLT-exact metrics measure strict string agreement with the coder-provided label but fail to capture clinically acceptable alternatives when multiple LLTs represent closely related concepts. To assess clinical acceptability for safety reporting, all RAG pipeline predictions underwent clinical expert review and were summarized using the Clinical Correctness Rate (CCR). Across datasets, clinical review indicated that a considerable fraction of apparent “errors” under LLT-exact evaluation were clinically acceptable alternatives (38-41%) (e.g., synonyms or near-equivalent LLTs appropriate for regulatory reporting). Clinical review yielded CCR values of 91-97% across datasets. These findings indicate that clinically meaningful performance cannot be fully captured by strict lexical agreement alone. Interrater agreement analysis showed moderate to substantial concordance across reviewers, with Fleiss’ kappa values ranging from 0.58 to 0.81 across datasets (See Supplement). In many cases, predicted LLTs differed from coder-assigned labels because they reflected closely related synonyms, alternative levels of granularity, or otherwise defensible regulatory phrasing. Thus, a substantial subset of apparent LLT-level “errors” arose within semantically dense regions of MedDRA where more than one LLT could plausibly represent the same AE narrative.

### Semantic Structure Explains Apparent LLT-Level Mismatches

To contextualize LLT-level disagreements, we analyzed the semantic structure of the LLT embedding space using Uniform Manifold Approximation and Projection (UMAP). As shown in Figure 3(a), LLTs belonging to the same System Organ Class (SOC) formed coherent clusters, indicating that the embedding model captures clinically meaningful relationships beyond surface lexical similarity.

**Figure 3.**
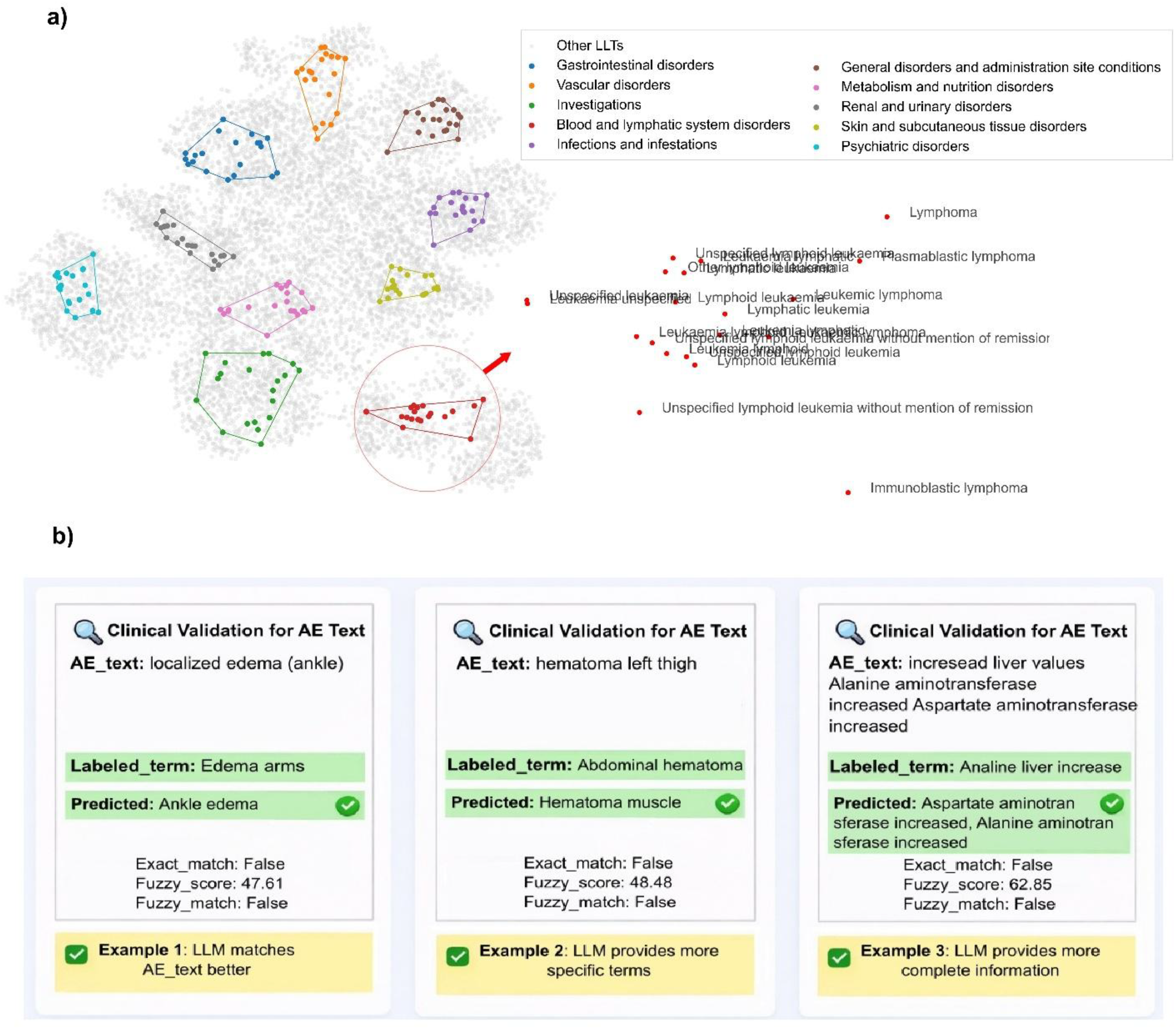
Semantic structure of the embedding space and clinical relevance of apparent LLT mismatches. a) Two-dimensional UMAP projection of MedDRA Low Level Term (LLT) embeddings (MiniLM), colored by primary System Organ Class (SOC). LLTs from the same SOC form coherent clusters, indicating preservation of clinically meaningful semantic structure beyond surface lexical similarity. A zoomed-in region highlights dense areas within the Blood and lymphatic system disorders SOC, where multiple closely related LLTs (e.g., leukemia- and lymphoma-related terms) occupy overlapping semantic space. b) Representative cases from clinical validation where model predictions differed from the human-baseline LLT but were judged clinically appropriate. These examples illustrate how LLT-level mismatches can arise from semantic ambiguity or differences in granularity rather than incorrect clinical interpretation.

**Figure 4.**
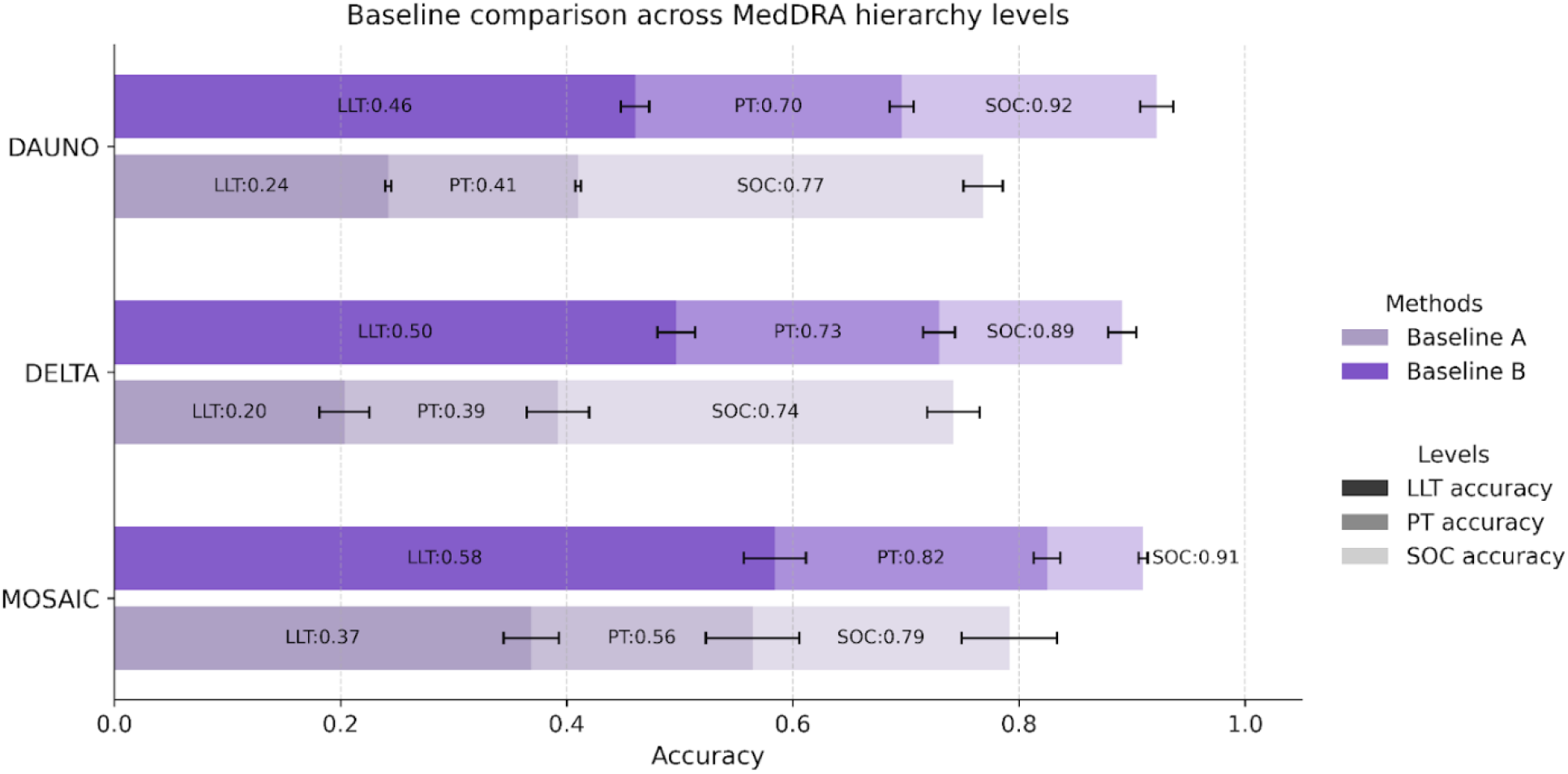
Performance comparison across MedDRA hierarchy levels under different candidate generation strategies. Accuracy at the LLT, PT, and SOC levels is shown for Baseline A (randomly sampled LLT candidates) and Baseline B (lexically retrieved candidates) across three AML clinical trial datasets. Bars denote mean accuracy over three random seeds. Error bars indicate standard deviation. **Abbreviations:** LLT = Low Level Term; PT = Preferred Term; SOC = System Organ Class; RAG = Retrieval-Augmented Generation; here used to refer to the RAG-inspired retrieval-augmented reasoning pipeline.

Zoomed-in regions reveal that certain SOCs (e.g., Blood and lymphatic system disorders) contain LLTs with very high semantic proximity. In such clusters, distinctions between closely related terms (e.g., lymphoid leukemia and lymphocytic leukemia) are subtle, and multiple LLTs may represent plausible interpretations of the same adverse event. Accordingly, some apparent LLT-level mismatches under automated evaluation reflect alternative but clinically acceptable coding choices rather than incorrect predictions. Representative examples of this phenomenon are shown in Figure 3(b).

### Semantic Candidate Retrieval Outperforms Random and Lexical Candidate Construction

We compared three approaches for selecting the most appropriate LLT from a candidate set. The randomly sampled candidate approach (Baseline A) achieved exact-match accuracy comparable to the zero-shot condition when evaluated against coder-provided reference labels (LLT exact: 20-37%, macro-F1 below 17%). Because the candidate list in Baseline A was generated by uninformed random sampling from the MedDRA vocabulary, exposure of the coder-assigned LLT was infrequent. This suggests that any successful selections in this condition were driven largely by the model’s internal knowledge rather than by informative candidate construction.

With Baseline B, we tested whether a candidate list generated based on lexical similarity performs comparably to the RAG approach, which retrieves semantically similar candidates. For example, “stomach ache” and “abdominal pain” are semantically similar but share little lexical overlap, whereas “abdominal pain” and “abdominal discomfort” exhibit higher lexical similarity due to their similar wording. Baseline B achieved LLT exact-match accuracy of 46-58% against the coder-provided reference labels. To directly quantify dense retrieval quality, we additionally assessed whether the coder-assigned LLT was present in the primary semantic top-100 candidate list before the reasoning step. Lexical fallback was used only in rare low-confidence or embedding-unavailable cases and therefore did not alter the interpretation that dense semantic retrieval served as the principal exposure mechanism. Dense retrieval exposed the coder-assigned LLT more reliably than lexical filtering, with inclusion rates of 88% in MOSAIC, 76% in DELTA, and 80% in DaunoDouble (overall approximately 81%), compared with 72%, 66%, and 66%, respectively, for Baseline B candidate construction. This finding indicates that the main advantage of semantic retrieval is not merely higher lexical overlap, but more reliable exposure of the coder-assigned LLTs under heterogeneous AE phrasing. Performance was stable across repeated runs, with low standard deviations across seeds for all reported metrics, indicating that the observed differences between candidate construction strategies were reproducible rather than driven by run-specific variation.

### Backbone Robustness Across Open Instruction-Tuned LLMs

To assess backbone sensitivity beyond the primary LLaMA-3.3-70B-Instruct configuration, we benchmarked the same retrieval-augmented reasoning pipeline across multiple open instruction-tuned LLMs using the MOSAIC, DELTA, and DaunoDouble AML trial datasets (Figure 5). Across models, agreement at the SOC level remained consistently high, and PT-level performance varied only modestly. The largest backbone-dependent differences were observed at the LLT level, where exact agreement requires finer discrimination among semantically proximate candidate terms. These findings suggest that the retrieval-augmented design maintains comparatively consistent higher-level MedDRA coding across different LLM backbones, while backbone choice mainly influences fine-grained LLT selection.

**Figure 5.**
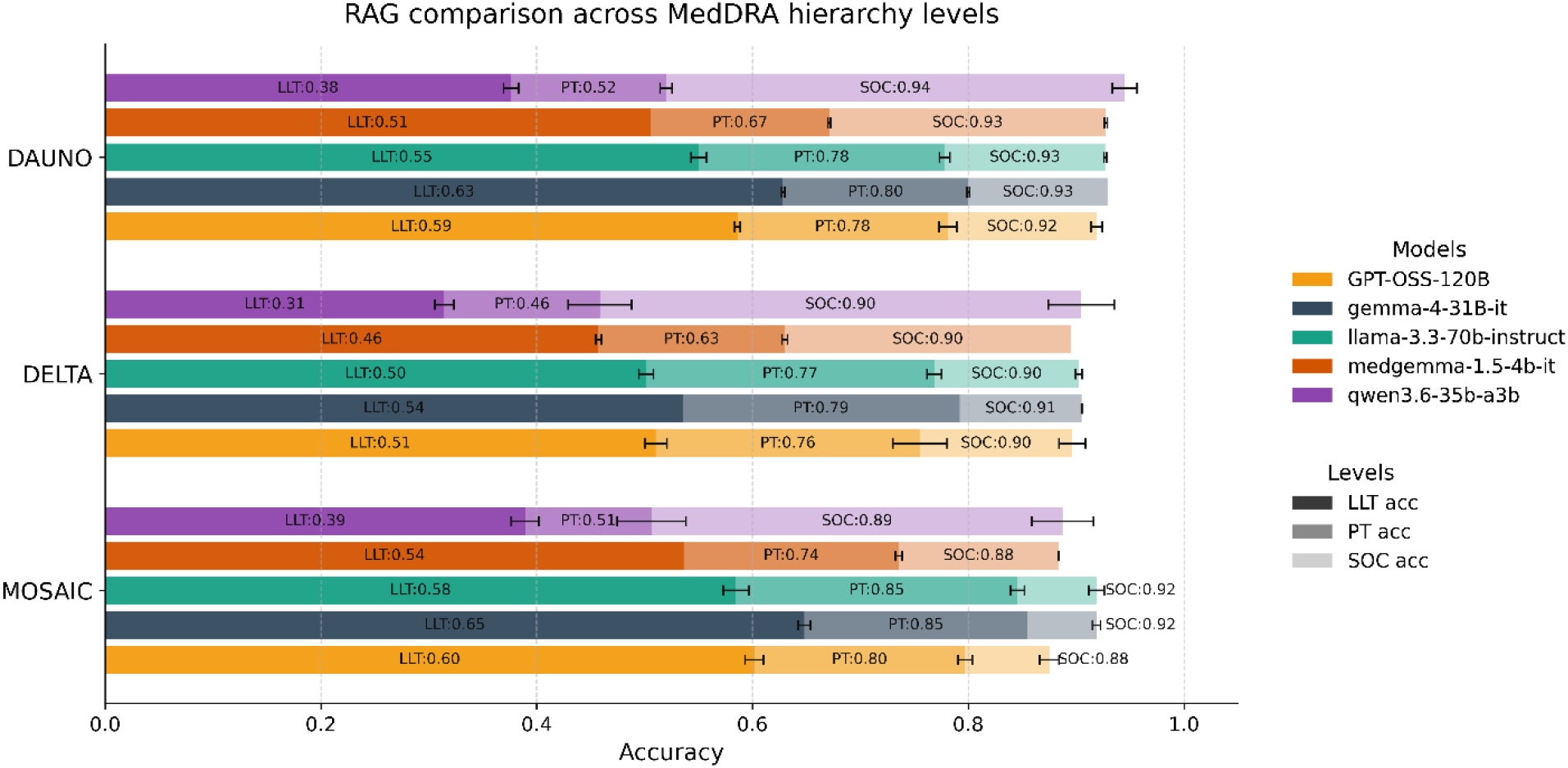
Multi-model benchmarking of the RAG pipeline across MedDRA hierarchy levels. Performance of the retrieval-augmented reasoning pipeline across three AML clinical trial datasets (MOSAIC, DELTA, and DaunoDouble) using five large language model backbones. Bars show mean LLT, PT, and SOC accuracies across three random seeds; error bars indicate standard deviation. Across models, SOC-level agreement remained consistently high, whereas the largest variation was observed at the LLT level, indicating that model choice primarily affects fine-grained LLT selection rather than higher-level MedDRA mapping. **Abbreviations:** LLT = Low Level Term; PT = Preferred Term; SOC = System Organ Class; RAG = Retrieval-Augmented Generation; here used to refer to the RAG-inspired retrieval-augmented reasoning pipeline.

## DISCUSSION

In this study, retrieval-augmented reasoning enabled clinically aligned MedDRA coding of freetext adverse events at the LLT level across three AML clinical trial datasets. The central finding is that fine-grained LLT-level coding becomes more reliable when dense semantic retrieval first narrows the terminology space to clinically plausible candidates and structured LLM reasoning then selects among semantically proximate alternatives. Retrieval and reasoning therefore served complementary roles: retrieval improved candidate exposure, while reasoning supported final selection among closely related LLTs. Importantly, the RAG pipeline should not be interpreted as a purely retrieval-driven improvement, because unlike the prompt-only baselines it also uses structured reasoning for final selection. Clinical expert review further showed that strict lexical agreement underestimated clinical appropriateness, as some LLT-level mismatches represented clinically acceptable alternatives rather than incorrect interpretations.

The poor performance of unguided zero-shot coding reinforces the need for explicit terminology grounding. Because zero-shot outputs rely on the model’s internal knowledge rather than a current MedDRA dictionary, they are less suitable for operational coding workflows where terminology versioning and traceability are essential. In this context, the RAG-LLM framework is better understood as a decision-support system for medical coders than as an unconstrained text-generation model.

Our findings are consistent with recent compositional and retrieval-augmented medical coding frameworks such as ALIGN and RuleAlign, which suggest that decomposing coding into constrained intermediate steps improves reliability over unguided generation (14,22). More broadly, work on LLM-assisted medical coding has emphasized that AI-assisted coding is most useful when embedded within structured clinical workflows rather than used as fully autonomous label-generation systems (7). Compared with previous work, our study contributes in three important respects. First, we operate directly at the LLT level, which is the most granular and operationally relevant level of the MedDRA hierarchy for trial documentation and regulatory reporting, whereas many prior approaches have focused on higher abstraction levels such as High-Level Terms (HLTs) or broader category-level coding (14,23). Second, we evaluate the system using adverse event narratives from three AML clinical trials rather than relying solely on historical coding benchmarks. Third, we complement automated lexical metrics with clinical expert review, allowing us to distinguish exact string disagreement from clinically acceptable alternative coding.

The use of three AML trial datasets provides a clinically relevant test setting for this framework. AML trials generate heterogeneous safety data across distinct treatment strategies and patient populations, while requiring consistent terminology for downstream safety interpretation and comparison (16,17). The high expert-assessed clinical correctness observed across MOSAIC, DELTA, and DaunoDouble therefore suggests that retrieval-augmented MedDRA coding may be particularly useful as a decision-support layer in hematology trial workflows, providing a framework for AI-supported pharmacovigilance.

Many existing AI-assisted coding systems rely heavily on lexical overlap or heuristic filtering (4,7,10), offer limited interpretability or lack explicit mechanisms for human-in-the-loop validation in safety-relevant settings (1,4,6). In contrast, the proposed framework supports transparency through explicit candidate lists, auditability through preserved predictions and deterministic LLT→PT→SOC mapping, and reproducibility through fixed prompts, deterministic decoding, and repeated seed-based evaluation. Although this does not constitute formal regulatory validation, these properties align with broader expectations for accountable AI in pharmacovigilance, including transparency, auditability, reproducibility, and expert oversight (6,22–24).

Notably, lexical filtering (Baseline B) and the RAG approach sometimes achieved similar LLT exact-match accuracy, but they should not be interpreted as equivalent. The RAG pipeline more reliably exposed the coder-assigned LLT within the candidate set, indicating that semantic retrieval provides a stronger foundation for accurate final selection than lexical filtering alone. In the implemented pipeline, dense semantic retrieval served as the primary candidate exposure mechanism, whereas lexical retrieval was reserved for rare fallback situations when dense retrieval was unavailable or did not meet predefined confidence criteria. Following clinical expert review, this distinction became even more apparent: several apparent LLT mismatches were judged clinically acceptable because they reflected semantically related terms, differences in granularity, or alternative but defensible regulatory phrasing rather than true coding errors. These findings highlight an inherent limitation of evaluating automated MedDRA coding solely by LLT exact-match accuracy. Because MedDRA LLTs include closely related synonyms, alternative levels of granularity, spelling variants, and other lexical variations, clinically appropriate predictions may still be penalized despite preserving the underlying clinical concept. Consequently, LLT exact accuracy should be interpreted together with PT/SOC-level metrics and clinical review to distinguish true clinical miscoding from acceptable terminology variation. These findings also help explain why earlier rule-based systems such as MedDRA Tagger (10), supervised adverse drug event extraction pipelines (4), and purely lexical pre-filtering methods remain vulnerable to synonymous, ambiguous, or institution-specific AE phrasing. Similar limitations have been reported for broader GPT-based medical text classification systems (11,25). In AML clinical trials, where heterogeneous adverse event descriptions are routinely compared across treatment arms and studies, this distinction between lexical disagreement and clinically acceptable coding alternatives is particularly relevant.

Taken together, our findings suggest that the RAG-LLM approach can support free-text AE-to-MedDRA LLT mapping in AML clinical trials, a task that remains labor-intensive and prone to inter-coder variability in routine coding practice (3,4,26). In this context, LLM-assisted systems should be viewed as decision-support tools that support more standardized MedDRA candidate selection while preserving expert oversight. Discrepancies between verbatim AE descriptions, coder-assigned LLTs, and hierarchical mappings further highlight the inherent variability of human coding decisions, echoing prior reports in pharmacovigilance and adverse event prediction (27). Prospective human-in-the-loop studies are needed to evaluate not only coding accuracy, but also coder efficiency, inter-coder consistency, user acceptance, and workflow integration in hematology clinical trial workflows.

Although our findings are encouraging, several limitations should be considered. This study was restricted to adverse event data from AML clinical trials. Broader validation across other hematologic malignancies, therapeutic areas, languages, institutions, and MedDRA versions will be important to establish the generalizability of the proposed framework. Automated evaluation also treated the coder-assigned LLT as the reference standard, although LLT-level MedDRA coding is granular and partly subjective. Several clinically acceptable model predictions differed from the human-assigned LLT and, in some cases, were judged at least as defensible as the reference label. Future studies should therefore evaluate equivalence or non-inferiority of LLM-derived LLTs in prospective blinded settings, for example by comparing human- and LLM-derived assignments side by side with independent adjudication. LLT-level coding errors can also propagate up the MedDRA hierarchy; therefore, direct PT-first prediction, uncertainty-triggered human review, and targeted adjudication of LLT/PT discordances deserve further investigation. Finally, although the system was evaluated retrospectively and in inference-only mode, multi-model benchmarking suggested that the retrieval-augmented design was comparatively robust across several open instruction-tuned LLMs, with the greatest variability observed at the LLT level and comparatively stable PT/SOC agreement. These future evaluations will be essential for determining the clinical utility and practical integration of LLM-assisted MedDRA coding into hematology clinical trial and pharmacovigilance workflows.

## CONCLUSION

Taken together, this work demonstrates that retrieval-augmented reasoning with large language models can support clinically aligned MedDRA coding of free-text adverse events in AML clinical trials by grounding LLT selection in explicitly retrieved MedDRA candidates. By focusing on end-to-end LLT-level prediction and combining automated evaluation with clinical expert review, our approach moves beyond surface-level lexical agreement and provides a practical foundation for future integration into hematology clinical trial and pharmacovigilance workflows through constrained candidate selection, deterministic hierarchical mapping, and structured expert validation.

Across three prospective AML clinical trial datasets, the proposed pipeline preserved strong PT/SOC-level agreement and achieved substantially higher expert-assessed clinical correctness than suggested by strict LLT-exact metrics alone. Multi-model benchmarking further indicated that the retrieval-augmented design was comparatively robust across open instruction-tuned LLM backbones, with the greatest variability remaining at the LLT level. These findings highlight the limitations of unguided generation and purely lexical candidate construction while supporting retrieval-augmented reasoning as a practical foundation for interpretable, human-in-the-loop decision-support systems for MedDRA coding in hematology clinical trials and pharmacovigilance settings.

## DATA AVAILABILITY

The datasets analyzed during the current study were derived from retrospective adverse event data from the MOSAIC (NCT04385290), DELTA (NCT02446145), and DaunoDouble (NCT02140242) clinical trials. Due to institutional, ethical, and patient privacy restrictions, the underlying clinical trial data are not publicly available. Aggregated results and methodological details supporting the findings of this study are included within the article and its Supplementary Information. Additional data may be made available from the corresponding author upon reasonable request and subject to institutional and regulatory approval.

## ETHICS STATEMENT

The study was conducted using retrospectively collected, fully de-identified adverse event data derived from three prospective clinical trials (MOSAIC, DELTA, and DaunoDouble). All original clinical trials were conducted in accordance with the Declaration of Helsinki and had received approval from the respective institutional ethics committees. This methodological study involved secondary analysis of anonymized data only and did not require additional participant recruitment or intervention.

## CONSENT

Not applicable, as the manuscript does not contain any individual person’s data in any form.

## CODE AVAILABILITY

The source code supporting this study will be made publicly available in a GitHub repository upon publication.

## AUTHOR CONTRIBUTIONS

N.D. conceived and designed the study, implemented the complete MedDRA-LLM pipeline, performed all computational experiments, developed the retrieval-augmented reasoning framework, conducted data analysis, created the prototype and interactive validation interface, prepared the figures, and wrote the main manuscript text. I.C.W. contributed to study supervision, manuscript writing, scientific interpretation, and critical revision of the manuscript. J.N.K. contributed to scientific interpretation, manuscript writing, and critical revision of the manuscript. M.M.K.S., J.-N.E., and M.M. contributed to clinical trial data acquisition and dataset preparation. I.C.W., M.M.K.S. and J.-N.E. additionally participated in clinical validation and expert review of model predictions. F.F., D.S., S.B., M.B., and C.R. contributed clinical expertise, reviewed the manuscript, and provided scientific feedback and comments. All authors reviewed, edited, and approved the final manuscript.

## ACKNOWLEDGEMENTS

The authors thank the clinical investigators, trial coordinators, and data management teams of the MOSAIC, DELTA, and DaunoDouble studies for their contributions to adverse event documentation and data curation. The authors further acknowledge the institutional computational infrastructure and technical support that enabled large-scale inference with large language models and bench-marking experiments.

