## Supplementary for "Retrieval-Augmented Large Language Models for Clinically Aligned Adverse Event Coding in Acute Myeloid Leukemia Clinical Trials"

### Lexical Fallback for Low-Confidence Dense Retrieval

Dense semantic retrieval served as the primary candidate generation strategy throughout the RAG-based pipeline. For each AE description, the top-k LLTs were first retrieved using cosine similarity over MiniLM embeddings. In rare cases where AE embeddings were unavailable or dense retrieval did not meet predefined retrieval-confidence criteria, lexical fallback retrieval using RapidFuzz (20) was applied to supplement candidate exposure. If the resulting candidate list still contained fewer than k items, it was completed with a small number of randomly sampled LLTs to preserve a fixed candidate list size. This fallback mechanism was intended to improve robustness without replacing dense retrieval as the principal retrieval layer. Embeddings were used exclusively for dense semantic candidate generation in the retrieval-augmented reasoning pipeline and were not used by any of the prompt-only baseline methods.

### Detailed Evaluation Metrics and Clinical Validation Protocol

Model performance was assessed using a combination of automated exact-match, lexical and hierarchical metrics together with clinical expert validation. This multi-level evaluation was designed to distinguish surface-level string agreement from clinically appropriate MedDRA coding.

*LLT Accuracy (Exact Match)*. Exact LLT accuracy was defined as the proportion of predictions in which the model-selected LLT exactly matched the coder-provided LLT label. This metric reflects strict lexical agreement with the coder-provided reference and does not capture clinical equivalence when alternative but clinically acceptable LLTs exist.

*Fuzzy LLT Accuracy*. To account for minor lexical variations, such as pluralization, spelling differences, or orthographic variants, fuzzy LLT accuracy was computed using lexical string similarity, with a similarity threshold of ≥0.90. This metric provides a more tolerant measure of lexical agreement than exact matching but does not assess clinical correctness.

*PT and SOC Accuracy (Hierarchical Evaluation)*. Because regulatory safety analyses are typically conducted at higher MedDRA levels, we additionally evaluated agreement at the Preferred Term (PT) and System Organ Class (SOC) levels. PT accuracy reflects agreement between the PT associated with the predicted LLT and the PT associated with the coder-provided LLT, whereas SOC accuracy reflects agreement at the primary SOC level. These hierarchical metrics provide broader measures of agreement when LLT-level lexical alignment differs.

*Precision, Recall, and Macro-F1*. To account for class imbalance across LLTs and PTs, we report precision, recall, and macro-averaged F1 scores for all methods and datasets.

*Clinical Correctness Rate (CCR)*. To directly assess clinical validity, all predictions generated by the retrieval-augmented reasoning pipeline underwent clinical expert review. Two independent clinicians evaluated whether each model-predicted LLT was clinically acceptable for regulatory reporting, irrespective of exact lexical agreement with the coder-provided label. Reviews were conducted independently and categorized as clinically acceptable or clinically incorrect. In cases of disagreement, a third senior clinician served as an independent adjudicator to establish a final consensus judgment. The Clinical Correctness Rate (CCR) was defined as the proportion of predictions deemed clinically acceptable under this consensus review.

*Inter-rater Agreement Analysis*. To quantify agreement among clinical reviewers, inter-rater agreement was calculated for the binary clinical validation labels (clinically acceptable vs. clinically incorrect). Fleiss' kappa was 0.81 for MOSAIC, 0.58 for DELTA, and 0.75 for DaunoDouble, indicating moderate to substantial agreement across datasets. Pairwise reviewer agreement was high, ranging from 98.3–99.4% in MOSAIC, 92.0–94.7% in DELTA, and 95.9–97.3% in DaunoDouble. These results indicate that the high Clinical Correctness Rate was not driven by a single reviewer and that reviewer disagreement was limited. The lower kappa observed in DELTA reflected greater reviewer variability in a subset of borderline cases: all three reviewers agreed on 203 of 226 cases, including 194 cases judged clinically acceptable and 9 cases judged clinically incorrect, while 23 cases showed partial disagreement. This pattern is consistent with the granularity and subjectivity of LLT-level MedDRA coding.

### Experimental Setup

Models were hosted on institutional GPU infrastructure and accessed via an OpenAI-compatible internal API; no commercial API usage or external billing was involved. The primary backbone was LLaMA-3.3-70B-Instruct, configured with temperature = 0.0, maximum generation length = 512 tokens, and context size = 128k. To assess backbone robustness, we additionally benchmarked the same retrieval-augmented reasoning pipeline across several alternative open instruction-tuned LLMs using the same retrieval, prompting, and evaluation protocol. Although decoding was deterministic (temperature = 0), repeated runs with different random seeds were used to capture variability introduced by candidate sampling and fallback mechanisms. Each experiment was repeated with random seeds {42, 43, 44}; we report mean ± standard deviation across runs. Dataset sizes are MOSAIC (n=179), DELTA (n=226), DAUNO (n=443); repeated runs do not change n but provide variance estimates.

**Table S1. Model hyperparameters and configurations.** Overview of institutional setup, backbone model, embedding and retrieval methods, prompting logic, and deterministic MedDRA mapping used in the MedDRA-LLM RAG pipeline.
**Abbreviations**: AE = Adverse Event; LLT = Low Level Term; PT = Preferred Term; SOC = System Organ Class; LLM = Large Language Model; API = Application Programming Interface; GPU = Graphics Processing Unit; MiniLM = all-MiniLM-L6-v2 sentence transformer; RAG = Retrieval-Augmented Generation; here used to refer to the RAG-inspired retrieval-augmented reasoning pipeline.

| Component | Configuration |
|---|---|
| **Institutional setup** | GPU server; models served via OpenAI-compatible API |
| **Primary LLM backbone** | LLaMA-3.3-70B-Instruct |
| **Decoding** | Deterministic; temperature = 0.0; maximum generation length = 512 tokens |
| **Context window** | → 128k tokens (prompts were ≤3.5k tokens in practice) |
| **Runs / reporting** | 3 repetitions per experiment; results as mean ± standard deviation |
| **Embedding model** | all-MiniLM-L6-v2, 384-dim sentence embeddings (LLT & AE) |
| **Retrieval (dense)** | Cosine similarity AE↔LLT; Top-K = 100 candidates |
| **Fallback retrieval** | In rare cases where AE embeddings were unavailable or dense retrieval did not meet predefined retrieval-confidence criteria, RapidFuzz (token-set similarity) was used as lexical fallback; if needed, random LLTs were appended to preserve a fixed candidate set size |

| | |
|---|---|
| **Prompting** | Structured retrieval-augmented reasoning with brief clinical justification before constrained LLT prediction: short reasoning → final line “Final answer: <LLT_TERM>” (robust parsing) |
| **Mapping** | Deterministic LLT → PT → SOC via MedDRA v25.0 lookups |
| **Primary SOC rule** | Priority: Ist_Primary_SOC = ‘Y’ → unique Primary_SOC_Code → single SOC → none |
| **Random seeds** | 42,43,44 (for fallback sampling) |

**Table S2. Performance of baseline methods and the RAG-based pipeline across MedDRA hierarchy levels**. Results are reported as mean ± standard deviation across three random seeds. Performance is shown for Baseline 0 (Zero shot), Baseline A (randomly sampled LLT candidates), Baseline B (lexically retrieved candidates via RapidFuzz), and the retrieval-augmented reasoning (RAG) pipeline, evaluated on three prospective AML clinical trial datasets (MOSAIC, DELTA, and DaunoDouble). Metrics include Low Level Term (LLT) accuracy under exact-match and fuzzy-match criteria (similarity ≥ 0.90), mapped Preferred Term (PT) accuracy, and System Organ Class (SOC) accuracy using hierarchical MedDRA mapping. Macro-averaged precision, recall, and F1-score are reported at the LLT level. Clinical validation metrics were assessed exclusively for the RAG pipeline. The Clinical Correctness Rate (CCR) reflects the proportion of predictions deemed clinically acceptable by clinical expert review, while the accepted error rate indicates the proportion of LLT-level exact-match errors that were judged clinically acceptable alternatives. SOC accuracies reported correspond to Option B (primary SOC of the mapped PT).

| Model / Dataset | LLT Accuracy (Exact) | LLT Accuracy (Fuzzy ≥0.90) | Mapped PT Accuracy | Mapped SOC Accuracy (Option B) | AE based SOC Accuracy (Option A) | Precision (macro) | Recall (macro) | F1 (macro) | Accepted Rate of Error LLT (%) | CCR (%) |
|---|---|---|---|---|---|---|---|---|---|---|
| **Baseline 0 – Zero-shot** | | | | | | | | | | |
| MOSAIC (*n=179) | 0.40 ± 0.00 | 0.51 ± 0.00 | 0.72 ± 0.00 | 0.74 ± 0.00 | 0.67 ± 0.01 | 0.23 ± 0.00 | 0.22 ± 0.00 | 0.22 ± 0.00 | – | – |
| DELTA (*n=226) | 0.28 ± 0.00 | 0.32 ± 0.02 | 0.59 ± 0.00 | 0.63 ± 0.00 | 0.57 ± 0.01 | 0.17 ± 0.00 | 0.17 ± 0.00 | 0.16 ± 0.00 | – | – |
| DAUNO (*n=443) | 0.37 ± 0.00 | 0.44 ± 0.00 | 0.63 ± 0.00 | 0.65 ± 0.00 | 0.60 ± 0.01 | 0.20 ± 0.00 | 0.22 ± 0.00 | 0.20 ± 0.00 | – | – |
| **Baseline A – Random Sampled LLT Candidates** | | | | | | | | | | |
| MOSAIC (*n=179) | 0.37 ± 0.02 | 0.42 ± 0.02 | 0.56 ± 0.04 | 0.79 ± 0.04 | 0.72 ± 0.06 | 0.19 ± 0.01 | 0.17 ± 0.01 | 0.17 ± 0.01 | – | – |
| DELTA (*n=226) | 0.20 ± 0.02 | 0.23 ± 0.02 | 0.39 ± 0.02 | 0.74 ± 0.02 | 0.62 ± 0.02 | 0.10 ± 0.01 | 0.10 ± 0.01 | 0.09 ± 0.01 | – | – |
| DAUNO (*n=443) | 0.24 ± 0.00 | 0.26 ± 0.00 | 0.41 ± 0.00 | 0.76 ± 0.01 | 0.70 ± 0.01 | 0.12 ± 0.00 | 0.11 ± 0.00 | 0.11 ± 0.00 | – | – |
| **Baseline B – Lexically Retrieved Candidates** | | | | | | | | | | |
| MOSAIC (*n=179) | 0.58 ± 0.02 | 0.67 ± 0.02 | 0.82 ± 0.01 | 0.90 ± 0.00 | 0.81 ± 0.00 | 0.42 ± 0.04 | 0.40 ± 0.04 | 0.40 ± 0.04 | – | – |
| DELTA (*n=226) | 0.49 ± 0.01 | 0.52 ± 0.01 | 0.72 ± 0.01 | 0.89 ± 0.01 | 0.77 ± 0.00 | 0.32 ± 0.02 | 0.33 ± 0.01 | 0.31 ± 0.02 | – | – |
| DAUNO (*n=443) | 0.46 ± 0.01 | 0.51 ± 0.00 | 0.69 ± 0.01 | 0.92 ± 0.01 | 0.85 ± 0.00 | 0.29 ± 0.02 | 0.30 ± 0.00 | 0.29 ± 0.00 | – | – |
| **RAG Pipeline** | | | | | | | | | | |
| MOSAIC (*n=179) | 0.58 ± 0.01 | 0.64 ± 0.00 | 0.85 ± 0.00 | 0.92 ± 0.00 | 0.85 ± 0.00 | 0.41 ± 0.01 | 0.37 ± 0.01 | 0.38 ± 0.01 | 0.39±0.01 | 0.97± 0.00 |
| DELTA (*n=226) | 0.50 ± 0.00 | 0.54 ± 0.00 | 0.78 ± 0.00 | 0.90 ± 0.00 | 0.85 ± 0.00 | 0.30 ± 0.01 | 0.30 ± 0.01 | 0.29 ± 0.01 | 0.41±0.02 | 0.91± 0.02 |
| DAUNO (*n=443) | 0.55 ± 0.00 | 0.60 ± 0.00 | 0.78 ± 0.00 | 0.93 ± 0.00 | 0.87 ± 0.00 | 0.36 ± 0.00 | 0.36 ± 0.00 | 0.35 ± 0.00 | 0.38±0.00 | 0.93± 0.00 |

* *Values are reported as mean ± standard deviation across three random seeds. n denotes the number of unique AE samples per dataset (not multiplied by runs). Note: An alternative SOC evaluation (Option A, AE-based SOC accuracy comparing coder-provided SOC with the predicted PT’s primary SOC) was computed for internal validation but was not used as a primary outcome measure in the main manuscript*.

### Prompt Templates

#### RAG pipeline prompt

```
f"You are a medical coding assistant. Your job is to reason through the best
MedDRA LLT term."
f"\nHere is an Adverse Event (AE):\n\"{ae_text}\"\n\n"
"Here is a list of candidate LLT terms:\n"
+ "\n".join(f"- {term}" for term in candidate_terms)
+ "\n\nPlease analyze the AE and list, and first provide a short reasoning."
"\nThen, on a separate line, write the best matching LLT in this format:"
"\nFinal answer: <LLT_TERM>"
```

#### Baseline A prompt

```
f"You are a medical coding assistant. Given the following adverse event
description:\n"
f"\"{ae_text}\"\n"
f"Choose the most appropriate MedDRA LLT term from the list below:\n\n"
+ "\n".join(f"- {term}" for term in sampled_terms)
```

```
+ "\n\nRespond only with the exact chosen term."
```

Baseline B prompt

```
f"You are a medical coding assistant helping to find the best matching MedDRA LLT term.\n"
f"Here is an adverse event description:\n\"{ae_text}\"\n"
f"Below is a list of possible MedDRA LLT terms. Select exactly one term that best fits the description.\n"
f"Respond only with the exact chosen term, without any extra text. \n\n"
+ "\n".join(f"- {term}" for term in closest_terms)+ "\n"
```

Zero-shot prompt

```
f"You are a medical coding assistant.\n"
"Your task is to assign ONE MedDRA Low Level Term (LLT) from the MedDRA dictionary" that best matches the following adverse event description. \n\n"
f"Adverse event description:\n\"{ae_text}\"\n\n"
"Respond with exactly one MedDRA LLT term, without any explanation, on a single line."
```

### Interactive User Interface for MedDRA Coding and Clinical Review

To illustrate the potential integration of the proposed framework into hematology clinical trial and pharmacovigilance workflows, we implemented a lightweight interactive user interface supporting both single-case inference and batch processing of adverse event (AE) data. The interface enables spreadsheet-based input of AE descriptions, automated MedDRA LLT prediction with hierarchical PT/SOC mapping using the retrieval-augmented reasoning pipeline, and structured human-in-the-loop validation. Users can review model outputs, select alternative candidate LLTs, manually override predictions, or accept and validate the suggested coding. Although intended as a research prototype, the interface demonstrates how AI-assisted MedDRA coding can be integrated with expert review in practical clinical coding workflows.

a) 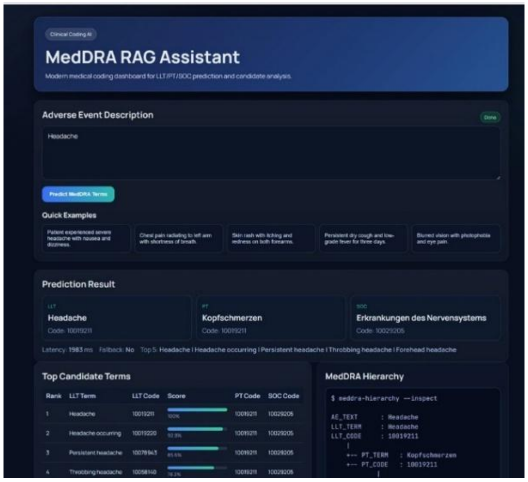


b) 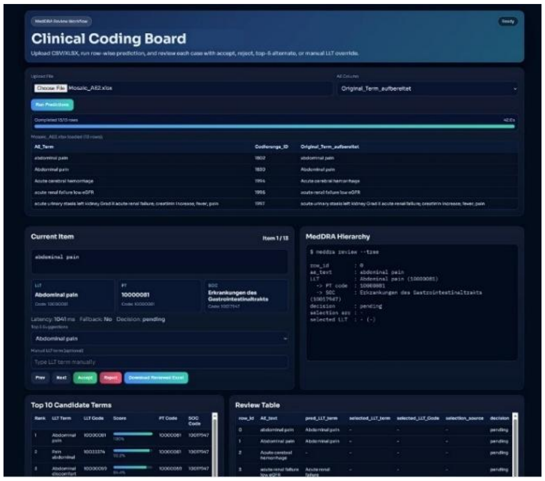


**Figure S1. Interactive user interfaces for MedDRA coding and human-in-the-loop validation**. a) Single-case inference interface for free-text AE-to-MedDRA coding. Users input a free-text adverse event description and receive predicted LLT, PT, and SOC assignments together with ranked candidate LLTs and hierarchical mapping. b) Batch-processing and review interface. Users can upload spreadsheet-based AE datasets, run row-wise MedDRA coding using the RAG pipeline, and perform structured clinical validation by accepting, rejecting, or correcting predicted LLTs, including selection from top candidate terms or manual override. The interface supports structured expert review and export of validated results.
